# Immortal time bias in older vs younger age groups: a simulation study with application to a population-based cohort of patients with colon cancer

**DOI:** 10.1101/2022.03.11.22272268

**Authors:** Sophie Pilleron, Camille Maringe, Eva JA Morris, Clémence Leyrat

## Abstract

**Background:** In observational studies, the risk of immortal time bias (ITB) increases with the likelihood of early death, itself increasing with age. We investigated how age impacts the magnitude of ITB when estimating the effect of surgery on one-year overall survival (OS) in patients with stage IV colon cancer aged 50-74 and 75-99 in England.

**Method:** Using simulations, we compared estimates from a time-fixed exposure model to three methods addressing ITB: time-varying exposure, delayed entry, and landmark methods. We then estimated the effect of surgery on one-year OS using a national population-based cohort of patients derived from the CORECT-R resource.

**Results:** In simulations, the magnitude of ITB was larger among older patients when their probability of early death increased or treatment was delayed. The bias was corrected using the methods addressing ITB. When applied to CORECT-R data, these methods yielded smaller effects than the time-fixed exposure approach but effects were similar in both age groups.

**Conclusion:** ITB must be addressed in all longitudinal studies, particularly, when investigating the effect of an exposure on an outcome in different groups of people (e.g., age groups) with different distributions of exposure and outcomes.

## 1. Introduction

Immortal-time bias occurs in longitudinal studies when the exposure is defined based on information available after the start of participants’ follow-up. This is typically the case when the start of follow-up and treatment initiation do not coincide^1^. In cancer literature, a classic example of ITB is when one wants to estimate the effectiveness of a treatment on survival by comparing survival measures (e.g. median survival, overall survival) from cancer diagnosis between patients who do, and do not, receive it. In practice, treatment is rarely initiated on the day of diagnosis, and therefore, in order to initiate the treatment at some point in time, patients must remain alive at least until the time of the treatment receipt; this period is, therefore, called “immortal time”. By defining the study groups based on the observed treatment assigned later, patients who may have been offered the treatment but died before initiation would contribute to the untreated group and as such inflate the number of deaths in that group. In a hypothetical trial in which patients are randomised to treatment groups at time of diagnosis, patients who die before treatment initiation would be on average equally represented in both study groups, and the ITB would not be a concern. In non-randomised study design, however, patients in the treated group would have an apparent survival advantage compared to those in the non-treated group, regardless of the efficacy of the treatment studied. Any apparent survival benefit in the treatment group may not, therefore, indicate a benefit of the treatment.

Several recent papers in epidemiology from different medical fields (oncology, nephrology, cardiology, etc.) drew attention to this bias^1–5^. However, it seems that this bias is still commonly misunderstood or overlooked in the cancer survival literature. Indeed, ITB was commonly seen in recent literature reviews^6 7–9^. However, several statistical methods are available to address the issue, including using the treatment as a time-varying exposure, the delayed entry approach or conditioning on a given survival time (landmark time)^4,10^.

Patients aged 75 years or older are underrepresented in randomised clinical trials, and observational studies are often used to study the effectiveness of treatment in terms of survival, with sometimes comparison between older patients and younger patients. Yet, the magnitude of ITB increases with the likelihood of early death, which itself increases as chronological age increases. Therefore, the magnitude of the bias may worsen with age. However, to our knowledge, no studies evaluated the impact of age on the magnitude of the ITB in cancer research and assessed the performance of standard and suitable analysis methods to account for this bias.

This study, therefore, aims to describe how the magnitude of ITB may differ in relation to age when age modifies the risk of death, the likelihood of receiving the treatment, or both. It also investigates the utility of three analytical techniques to account for this bias in practice. Initially a simulation study was conducted to empirically illustrate the impact of age on the magnitude of ITB under different scenarios when using a time-fixed exposure statistical approach prone to ITB, and to compare the performance of three alternative methods to account for this bias. Then, these methods were applied to estimate the effect of surgery performed within 6 months of diagnosis on one-year overall survival in patients diagnosed with stage IV colon cancer aged 50-74 and separately, in those aged 75-99 in England using data from the CORECT-R resource^11^.

## 2. The problem

To estimate the effect of surgery performed within 6 months of diagnosis on the one-year overall survival probability from cancer diagnosis in those who do, and do not, receive the treatment and by age group one can use the Kaplan Meier estimator or a Cox regression model by simply including surgery status in the model, that is, whether the patients received surgery in the 6 months following the diagnosis or not. This considers treatment as a time-fixed exposure. These methods wrongly assume that surgery occurs at cancer diagnosis whereas in practice, this may not be the case.

To illustrate the risk of ITB obtained using standard statistical approaches, we generated simplified data based on our illustrative example, for the estimation of the effect of surgery within 6 months on one-year overall survival in patients diagnosed with stage IV colon cancer aged 50-74 (called younger patients hereafter) and separately, in those aged 75-99 (older patients hereafter). Data were generated under four scenarios in which the probability and timing of death and the probability and timing of treatment could differ between age groups. For each scenario, we generated data following Weibull distributions for 100,000 patients, 50,000 in the younger group, and 50,000 in the older group (details below). For illustrative purposes, the data were generated under the hypothesis of no treatment effect. Then, for each patient, the surgical treatment status and time to surgery, and vital status and survival time were generated from Weibull distributions, with chosen parameters to correspond to the following scenarios:

– Scenario 1: younger and older patients have the same survival and treatment distributions.
– Scenario 2: the probability of having surgery within 6 months and the distribution of time to surgery, as well as the one-year survival probabilities are the same for the two age groups, but deaths occurred earlier among older patients.
– Scenario 3: the one-year survival probabilities and survival times, as well as the probability of having received surgery at 6 months are the same in the two age groups, but older patients are treated later on average.
– Scenario 4: the one-year survival probability and the probability of receiving surgery within 6 months are lower among older patients than among younger patients, and older patients die sooner and receive treatment later than younger patients.

The generated treatment status and time to treatment correspond to the intention to treat, that is, to what would have been observed if the patients could not die. However, for patients whose generated survival time was shorter than their generated time to surgery, their observed treatment status was set to 0. These scenarios are depicted in Figure 1.

**Figure 1.**
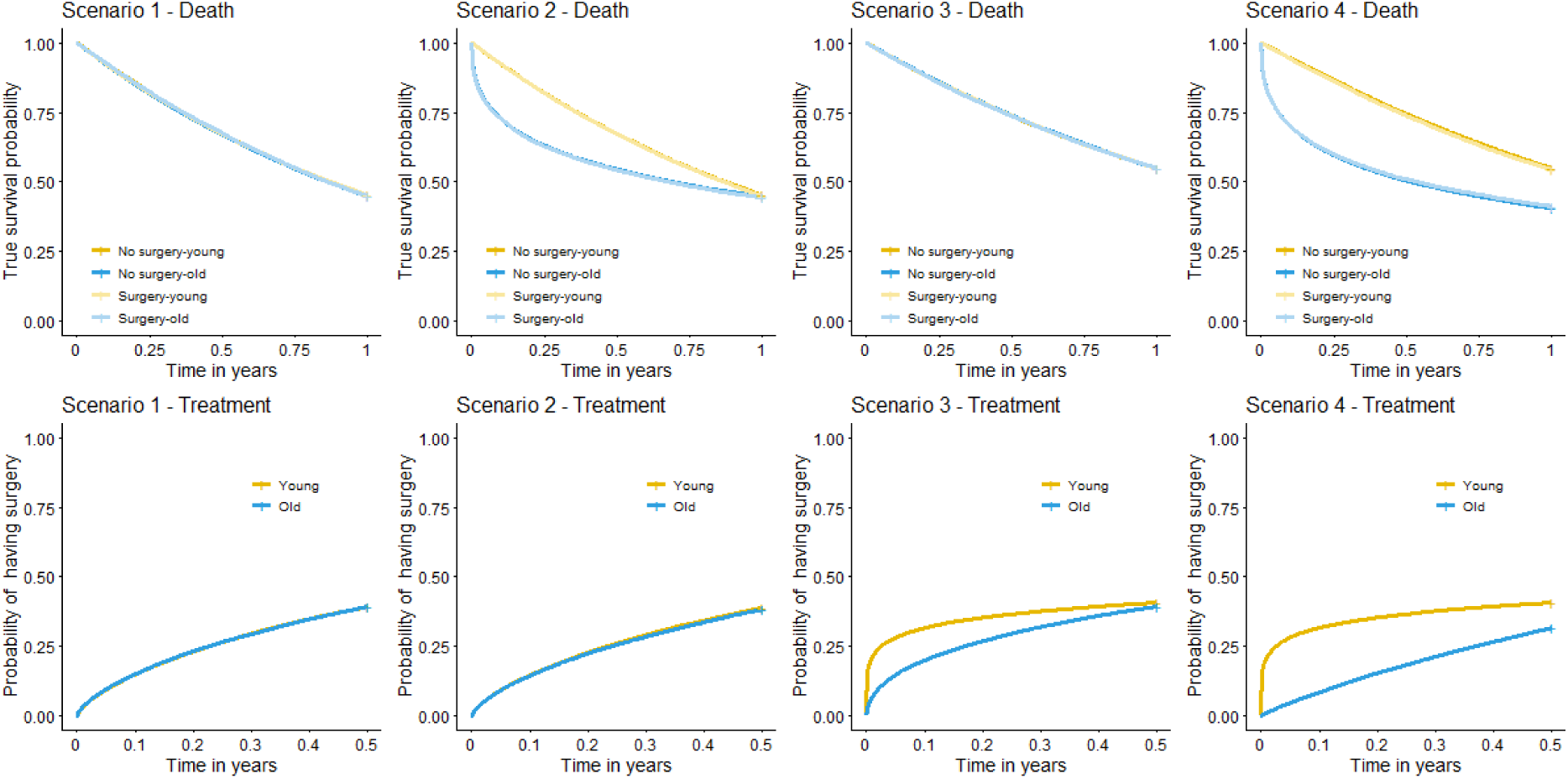
– Scenarios used in simulations

Then one-year overall survival in both treatment groups was estimated separately for younger and older patients, using a Cox regression model including treatment as a time-fixed exposure (Figure 2).

**Figure 2.**
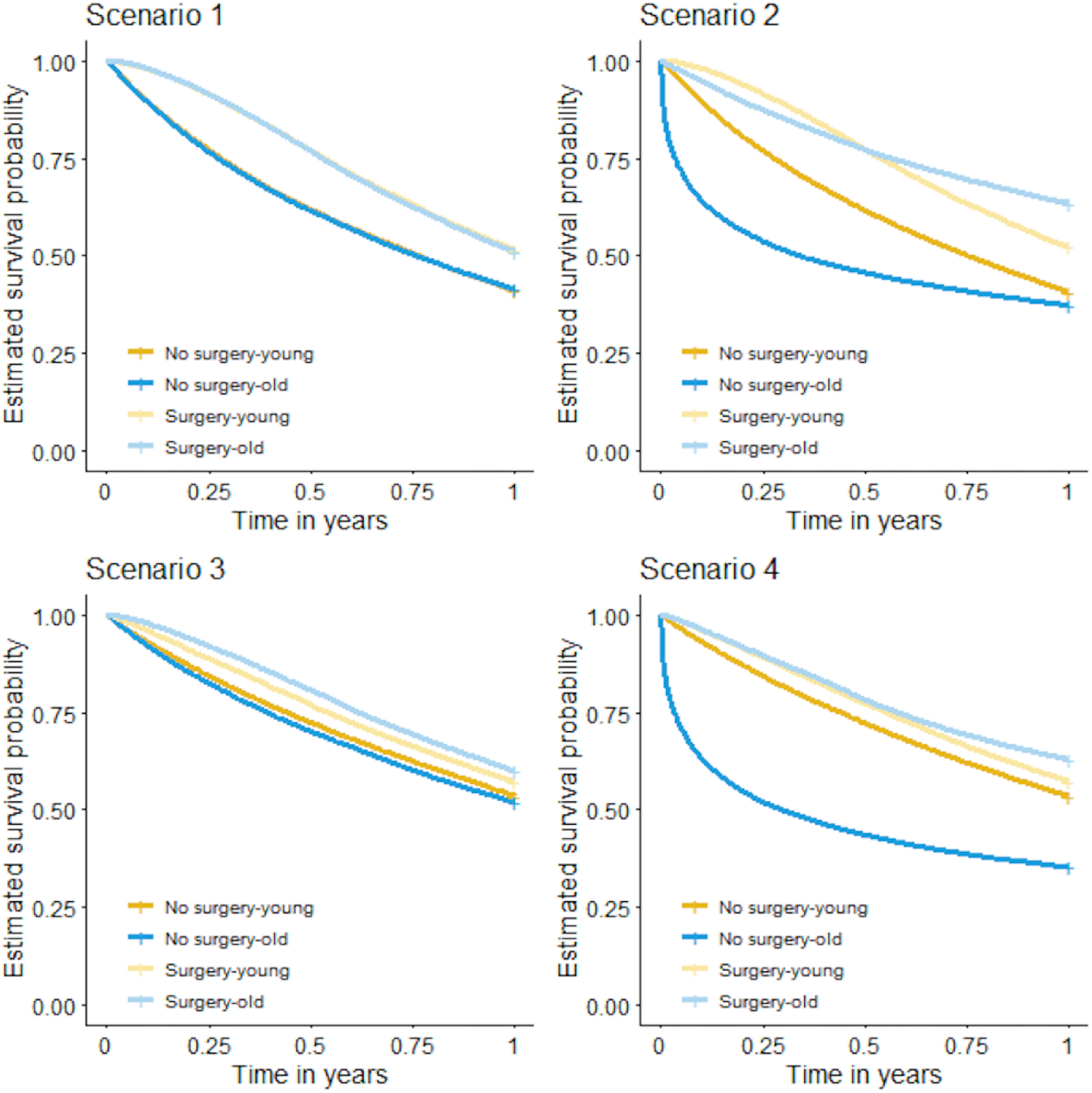
– Overall survival estimated using the time-fixed exposure method based on the four scenarios in young and old groups

In Scenario 1, surgery as a time-fixed treatment led to a substantial bias due to immortal-time: while the true treatment effect is 0 in both age groups, the observed differences in one-year survival probabilities were about 10% in both age groups. However, because survival and treatment distributions were identical among younger and older patients, the magnitude of the ITB was the same in the two groups. In scenarios 2 and 3, the magnitude of ITB was larger among older patients. In both cases, this is because the number of patients who died before receiving surgery was larger among older patients. This phenomenon was further amplified in scenario 4, in which older patients had a higher probability of death (Figure 2).

This illustrates the need for appropriate statistical analyses to obtain treatment effect estimates not affected by immortal-time bias.

## 3. Statistical methods for handling ITB

Several statistical methods have been proposed to address the ITB. One of them is to move forward the time 0 and setting up a new time 0 (landmark time) at a time where, for instance, most patients are likely to have received treatment, such as six months after diagnosis (researchers should define this period *a priori* for their research question based on clinical and field knowledge rather than data)^12,13^. All patients who died before the landmark time are excluded from the analysis. All patients who do receive treatment after the landmark time are categorized in the untreated group. This method is known as landmark analysis. It addresses the issue of ITB by conditioning on the survival time. The overall survival from diagnosis but among patients who survived at least until the landmark time can then be estimated using standard statistical methods, such as using Kaplan Meier estimator or a Cox regression model. Because of the exclusion of all deaths occurring before the landmark time, this method estimates the effect of treatment in patients who were alive at the landmark time, therefore this method estimates a conditional effect, and it cannot be interpreted as the marginal effect of surgery in the population.

Another technique considers patients as untreated until they are treated and treated thereafter. The patient treatment status therefore changes over time. This method is called time-varying exposure analysis^14^. Time-varying exposure analysis, such as the Cox model with treatment as a time-varying variable, can be used for analysis. Contrary to the landmark analysis, the whole sample is analysed, which allows the estimation of the marginal treatment effect from diagnosis on the entire population.

A third technique to handle immortal time bias is to allow for delayed entry. This method is essentially similar to the time-varying exposure method described previously, except that two models estimate separately the survival probabilities among the treated and the untreated. The model under no treatment includes the entire follow-up of the untreated patients, and the time between the diagnosis and the treatment for the treated patients. The model under treatment includes patients from the time of treatment to the end of follow-up. By having two models, this approach would be equivalent to the time-varying exposure approach in which all the covariates-treatment interactions would be included. However, a drawback is that unlike the time-varying exposure and landmark analyses, model-based standard errors cannot be obtained because there are two independent models involved, therefore, standard errors must be obtained using non-parametric bootstrap.

## 4. Simulation study

### 4.1 Method

#### Aims

The aim of the simulation study was to investigate under which circumstances the magnitude of the ITB differs between younger and older populations and to illustrate the performance of the time-varying approach, the delayed entry method and the landmark analysis for the estimation of treatment effects in the presence of ITB. The simulation study was conducted, therefore, to investigate the impact of age on ITB when no other biases are in play. This simulation is illustrative and does not aim to fully evaluate the statistical properties of the aforementioned methods.

#### Data generation

The data were generated as described previously in Section 2, except that the sample size was 1000 for each generated dataset. We considered 7 scenarios: the 4 scenarios introduced in Section 2, and the following 3 additional scenarios (see Figure 1 in Supplementary material):

Scenario 5: the probability of having surgery within 6 months and the distribution of time to surgery are the same for the two age groups, but deaths occurred earlier among older patients, and their one-year OS is lower.

Scenario 6: the one-year survival probabilities and survival times are the same in the two age groups, but older patients have a lower probability of treatment and are treated later on average.

Scenario 7 : the one-year survival probabilities and probabilities of treatment are the same in the two age groups, but older patients are treated later on average and die earlier.

Table 1 of the Supplementary material shows the values of the shape and scale parameters of the Weibull distributions in each scenario. In the 7 scenarios, no treatment effect in either age group was assumed. All the simulations were conducted in R 3.6.0^15^. We used the R package *simsurv* to generate the time-to-event data^16^.

**Table 1.** Characteristics of our study sample by age group.

|  | 50-74 years old | 75-99 years old |
| --- | --- | --- |
| n | 8389 | 8430 |
| Number of deaths within the first year after diagnosis (%) | 4311 (51.4) | 6530 (77.5) |
| Number of deaths within the first six months after diagnosis (%) | 3127 (32.3) | 5242 (62.2) |
| Median follow-up time in days (IQR) | 344 (81-365) | 106 (35-322) |
| Surgery - n (%) | 3777 (45.0) | 1994 (23.7) |
| Surgery within 6 months – n (% of all patients with surgery) | 3435 (90.9) | 1940 (97.3) |
| Median age in years (IQR) | 66 (60-70) | 82 (78-86) |
| Females - n (%) | 3799 (45.3) | 4297 (51.0) |
| Deprivation index quintiles – n (%) |  |  |
| 1 – Less deprived | 1776 (21.2) | 1830 (21.7) |
| 2 | 1861 (22.2) | 1927 (22.9) |
| 3 | 1677 (20.0) | 1848 (21.9) |
| 4 | 1579 (18.8) | 1559 (18.5) |
| 5 – Most deprived | 1496 (17.8) | 1266 (15.0) |
| Charlson's comorbidity index – n (%) |  |  |
| 0 | 6115 (72.9) | 4467 (53.0) |
| 1-2 | 1757 (20.9) | 2657 (31.5) |
| 3+ | 517 (6.2) | 1306 (15.5) |

IQR: Interquartile range

#### Estimands

Our estimand of interest was the difference in one-year survival probabilities following diagnosis between treated and untreated patients, among younger and older patients separately.

#### Methods

In each generated dataset, we estimated the difference in one-year overall survival probabilities using a Cox regression model based on the time-fixed exposure approach as well as the three approaches addressing the ITB, which are landmark analysis, time-varying exposure analysis, and delayed entry method. The models were estimated separately by age group. For the time-varying Cox model, the data were split at each time a death occurs, meaning that a row was one patient per time interval. Non-parametric bootstrap was used to estimate the 95% confidence intervals for the difference in survival probabilities. Normal-based confidence intervals were constructed using 5000 replications.

#### Performance measure

We estimated the bias of the estimate of the difference in one-year survival probabilities, as well as the average bootstrap standard errors across simulations and empirical standard error.

### 4.2 Results

The results of the seven main scenarios are presented in Figure 3. As already illustrated in Section 2, using a time-fixed exposure leads to a strong bias in all the scenarios (bias ranging from -0.092 in scenario 3 to -0.319 in scenario 4). As expected, if there are no differences in terms of survival and probability and timing of treatment between age groups, the magnitude of the ITB is similar in the two age groups (Scenario 1). However, the magnitude of the ITB increases with earlier death and delayed treatment (scenarios 2 to 7), which are likely to be more common among older patients in comparison to younger patients. With a time-varying exposure, a delayed entry model and landmark analysis, the bias was corrected. None of these approaches led to convergence issues (convergence rate of 100% across scenarios). Out of the three unbiased approaches, the delayed entry methods led to the smallest empirical standard error across simulations (Table 2 in supplemental material).

**Figure 3.**
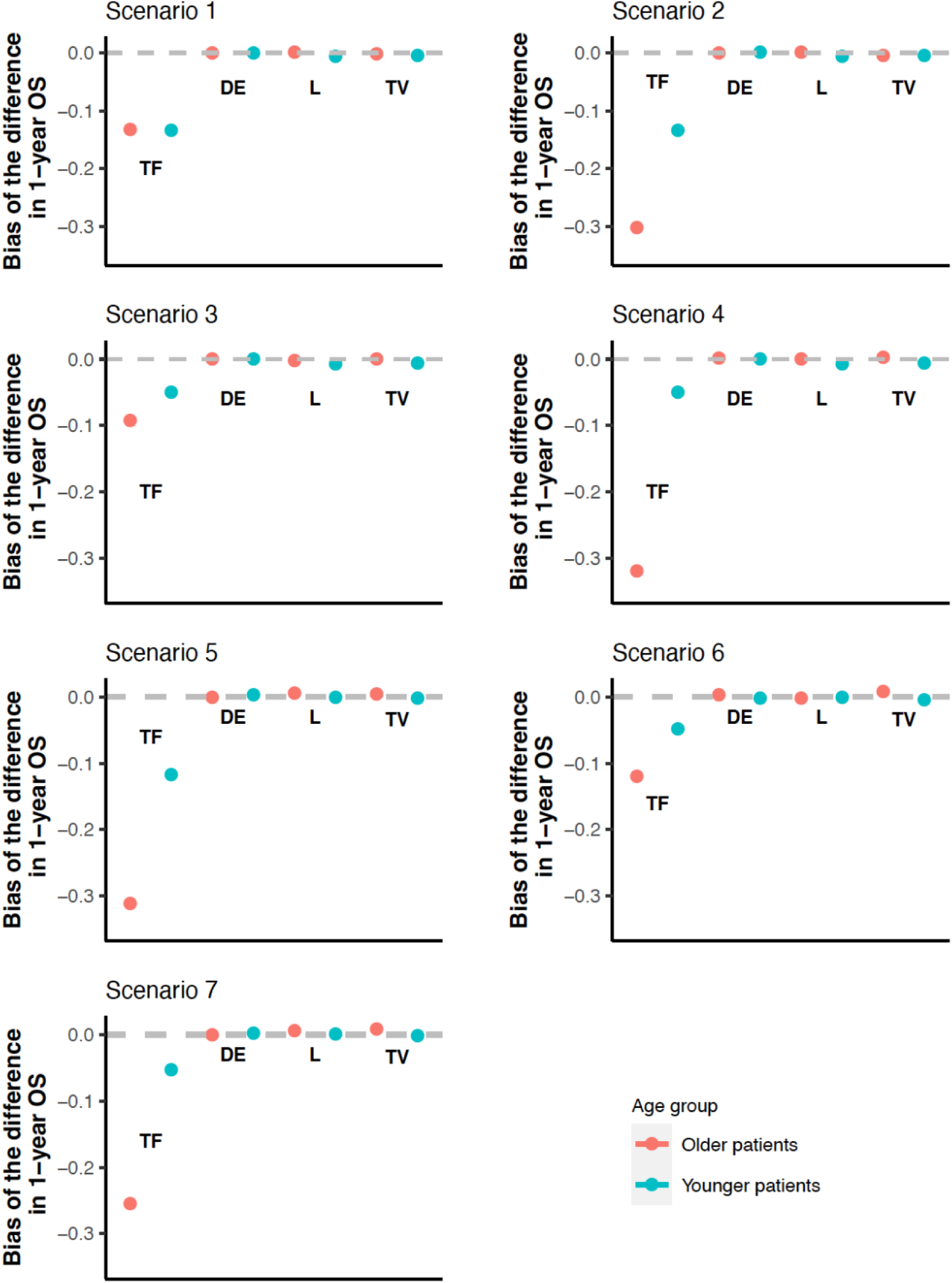
Bias of the difference in one year overall survival (OS) based on the seven scenarios TF: Time-fixed exposure method; DE: Delayed entry method; L: landmark method; TV: time-varying exposure method Note: a difference of 0 indicates there is no immortal time bias.

**Table 2.** One-year overall survival (%) by age category, receipt of surgery and method to address immortal time bias (reference group: no surgery group)

| Method | Estimand | 50-74 years old |  |  |  |  | 75-99 years old |  |  |  |  |
| --- | --- | --- | --- | --- | --- | --- | --- | --- | --- | --- | --- |
|  |  | No surgery |  | Surgery |  | Difference in 1-year survival | No surgery |  | Surgery |  | Difference in 1-year survival |
|  |  | n | One-year OS (95% confidence interval) | n | One-year OS (95% confidence interval) | - | n | One-year OS (95% confidence interval) | n | One-year OS (95% confidence interval) | - |
| Time-fixed exposure method | 1-year overall survival | 4954 | 31.9 (30.6; 33.2) | 3435 | 71.8 (70.3; 73.3) | **-39.9 (-41.8; -38.0)** | 6490 | 13.1 (12.2; 14.0) | 1940 | 52.3 (50.1; 54.5) | **-39.1 (-41.4; -36.8)** |
| Landmark analysis | 1-year overall survival conditioning on being alive beyond landmark time (i.e., 6 months) | 2326 | 69.6 (67.7; 71.5) | 2926 | 84.0 (82.7; 85.3) | **-14.4 (-16.7; -12.1)** | 1762 | 49.7 (47.3; 52.1) | 1415 | 72.7 (70.3; 75.1) | **-22.9 (-26.2; -19.6)** |
| Time-varying exposure | 1-year overall survival | 4954 | 36.0 (34.6; 37.4) | 3435 | 68.3 (66.7; 69.9) | **-32.4 (-34.4; -30.4)** | 6490 | 14.7 (13.8; 15.6) | 1940 | 47.7 (45.4; 50.0) | **-33.0 (-35.5; -30.5)** |
| Delayed entry method | 1-year overall survival | 4954 | 36.9 (35.5; 38.3) | 3435 | 68.3 (66.6; 70.0) | **-31.4 (-33.6; -29.2)** | 6490 | 15.1 (14.2; 16.0) | 1940 | 46.8 (44.1; 49.5) | **-31.7 (-34.5; -28.9)** |

## 5. Illustrative example

### 5.1 Data and Method

We included 10,392 patients aged 50-74 and 10,071 patients aged 75-99 years old diagnosed with stage IV colon cancer (ICD-10 code: C18) in England between 2014 and 2017 from the COloRECTal cancer Repository (CORECT-R). The CORECT-R is a national population-based resource that provide information about all patients diagnosed with colorectal cancer in England thanks to the linkage of cancer registry data to a variety of datasets including hospitalisation data from Hospital Episode Statistics (HES)^11^. We sequentially excluded patients with unknown vital status (n=16), unknown survival time (n=17), patients with no record in HES (n=419), and those whose surgery was anterior to the cancer diagnosis (n = 435). As we are interested in assessing the effectiveness of major resection against no surgery, we further excluded patients who had minor resection, stoma, stent, and bypass (n=2757), leaving 16819 patients for analysis. All patients were followed up to their death, or December 31, 2018, whichever comes first. We censored all patients alive beyond one year after diagnosis. Patients who underwent surgery later than six months after diagnosis were considered untreated. We estimated one-year OS in both age groups (i.e., 50-74 and 75-99) by surgery status (yes/no) using the time-fixed exposure method and the three methods accounting for ITB introduced previously. For the landmark analysis, we chose a landmark time at six months after diagnosis, most surgeries occurring within this time window. We excluded all patients who died before the landmark time. For the time-varying exposure approach, we ran a Cox regression model including treatment as a time-varying variable. For the delayed entry method, we fitted two separate Cox regression models in all untreated patients (including patients treated but censored at time of surgery) and treated patients, respectively.

All models were adjusted for age (restricted cubic spline with 1 interior knot placed at the median of the observed distribution of age, and two boundary knots placed at 10% and 90% quantile of the observed distribution of age), sex, quintile of socio-economic deprivation level and Charlson’s comorbidity index categorised into 0,1-2, 3+. We then estimated the marginal overall survival in the treated and untreated groups by predicting the individual one-year survival for all patients under two hypothetical scenarios: as if all the population was treated and then as if the whole population was untreated. The effect of surgery was the difference in mean one-year overall survival in these two hypothetical scenarios. Non-parametric bootstrap, using 5000 replications was used to calculate standard errors and derive 95% normal-based confidence intervals.

We performed statistical analyses using R statistical software (version 3.4.0; R Development Core Team, 2017). The R codes for the simulation study and the CORECT-R analysis are available in supplemental files 2 and 3, respectively.

### 5.2 Results

Table 1 presents patients’ characteristics by age group. Median follow-up time was shorter in older patients (106 days) as compared to younger patients (344 days). About a quarter of older patients underwent surgery against 45% of younger patients. A vast majority of surgeries occurred within 6 months from diagnosis in both groups. There were more women in the older age group, and older patients had a higher Charlson’s comorbidity index than younger patients. Deprivation level was similarly distributed in both age groups.

Figure 4 shows the observed distribution of time-to-death and time-to-treatment. In the first graph, older patients die sooner and have poorer one-year overall survival than younger patients in both treatment group. However, this representation does not take into account immortal time bias, precluding the direct comparison between curves. In the 2^nd^ graph, among patients who underwent surgery, older patients received surgery sooner than younger patients, however, the graph does not take into account higher mortality in older adults.

**Figure 4.**
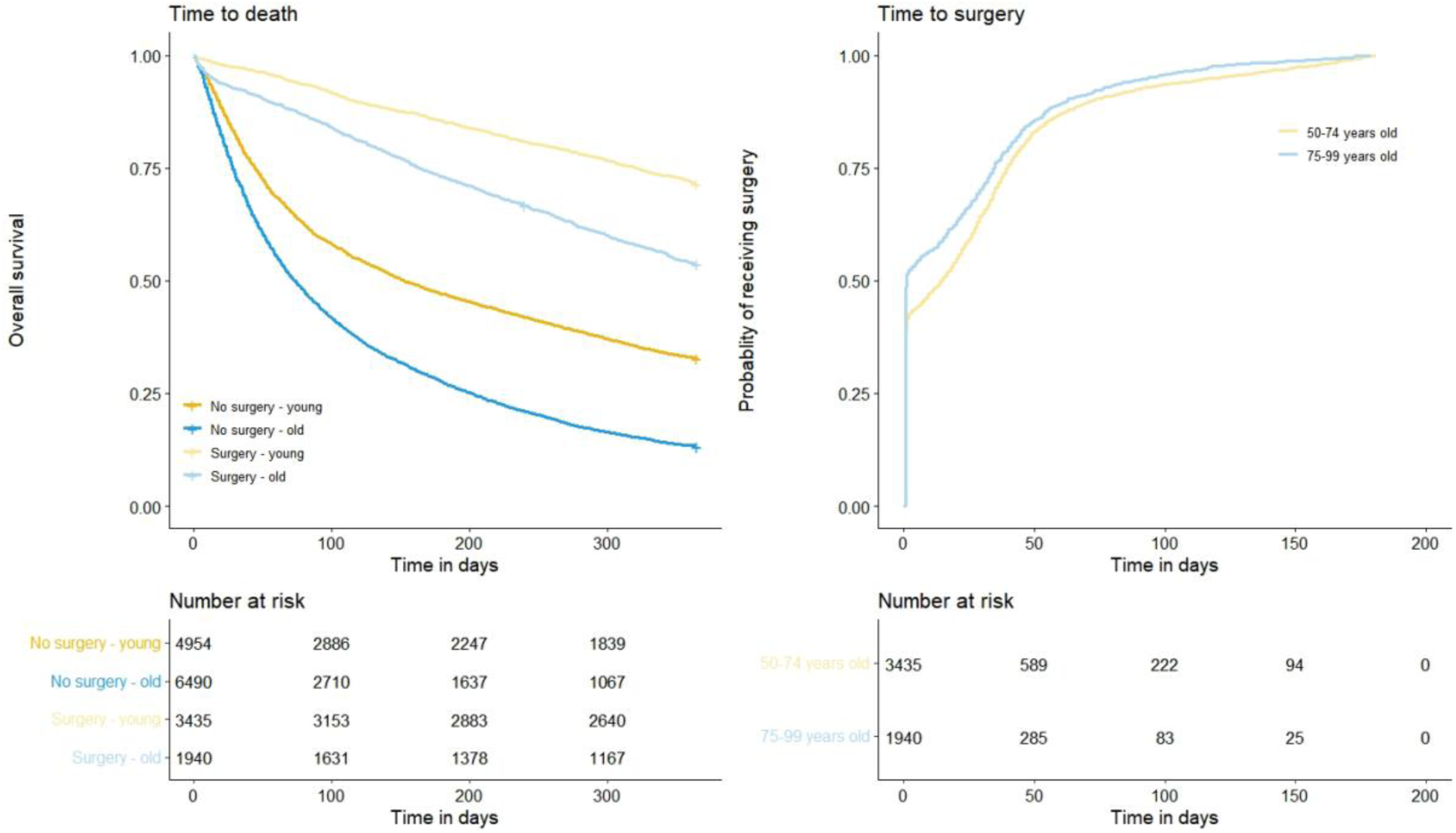
Distribution of time-to-death (i.e. survival time) and time-to-surgery in patients with stage IV colon cancer aged 50-74 and 75-99 who received surgery within 6 months of diagnosis and those who did not.

Table 2 presents one-year OS estimates in each treatment group and the difference in survival between the two treatment groups by age category and analysis method.

When compared to both the time-varying exposure and the delayed entry approaches, the time-fixed exposure approach led to a larger effect of surgery on one-year OS, than other methods, as expected. Both the time-varying exposure and delayed entry methods provided similar estimates of the effect of surgery on one-year OS in both age groups (about 20 percentage points). Besides, the difference in the effect of surgery between time-fixed exposure approach and the time-varying exposure or delayed entry method is about 8-9 points suggesting no difference in the magnitude of the ITB across age groups.

Using the landmark analysis, survival estimates were higher than those estimated using the other methods due to the exclusion of patients who died within the first six months after diagnosis. Given that a different sample is used for this analysis, a direct comparison of surgery effect estimates with other approaches is not possible. In patients who survived at least six months, the effect of surgery on the conditional one-year OS was higher in the 75-99 age group (−17 point difference; 95% Confidence Interval: -20;-13) than in the 50-74 age group (−10 point difference: - 12;-7).

## 6. Discussion

In a context where older patients with cancer are still less likely to be included in randomised clinical trials^17^, and more generally where randomised clinical trials are deemed unfeasible or unethical in some circumstances (e.g. resection of colorectal cancer liver and lung metastases^18^), population-based observational data (e.g. cancer registry, hospital statistics) are a good source of information that may be used for comparative effectiveness research. However, observational studies are prone to a certain number of biases, including ITB, and researchers have to be careful when planning their study. We show the magnitude of the ITB varies based on the distribution of death and patterns of treatment receipts over time. We provide R scripts for relevant statistical methods that corrects for the ITB.

We presented three methods that were proposed to control for the ITB: the landmark analysis, the analysis considering treatment as a time-varying exposure, and the delayed entry method. The first two methods are the most commonly used in the literature. Both the time-varying exposure and the delayed entry method estimate the effect of treatment on overall survival from cancer diagnosis (such as surgery on one-year survival in our study), while the landmark analysis estimates survival probabilities conditionally to surviving beyond a pre-defined landmark time (i.e., six months in our study). Consequently, the resulting point estimates cannot be compared with the two other approaches. The quantity to estimate will depend on the question asked: are we interested in the effect of surgery on survival beyond one year from diagnosis or are we interested in knowing the effect of surgery beyond one year since diagnosis if patients survive for at least a certain period of time after diagnosis, for instance because of high early mortality prior to treatment?

The landmark analysis is easy to implement and to interpret, but this method excludes patients who died before the landmark time. Therefore, the results depend on the choice of the landmark time. This method may provide useful information about survival prognosis for the clinical management of patients. Time-varying exposure analysis and the delayed entry method use the whole sample of patients for analysis. Delayed entry method is similar to the time-varying exposure method with the difference this approach requests to model the estimator of interest separately in the treated and the untreated groups. Therefore, it allows the effect of covariates to differ by treatment group.

Another way to handle ITB is to emulate a target clinical trial one would have conducted to answer the research question^19^. This method mimics a clinical trial using observational data, by explicitly defining time zero (i.e., time when patient meets eligibility criteria and is assigned to treatment group), the selection of patients, exposures, outcomes and the effect to estimate. The method relies on patients’ cloning, censoring and weighting to address ITB. However, the emulated trial estimates the intention-to-treat effect while the methods presented estimate the effect of surgery once received.

In our illustrative example, the effect of surgery was similar across age groups and across methods (excluding landmark analysis) suggesting the magnitude of ITB was similar across group (same difference between estimates obtained using fixed-time exposure method and varying-time exposure or delayed entry methods across age groups). This contrasts with the results of our simulation study which showed how the probability of receiving surgery, and the probability of death influenced the magnitude of the ITB. One hypothesis is that older patients may die sooner and received surgery earlier than younger patients, cancelling the respective effects on ITB. Because we used observational data, we cannot rule out residual confounding due to unmeasured confounding factors. Indeed, we regret the lack of information about patients’ fitness, geriatric conditions, social support, or care providers which play the role of confounder in the relationship between surgery and survival. Also, we did not make the distinction between emergency surgery and elective surgery. Older patients are more likely to be diagnosed through emergency presentation which is associated with poor survival prospects^20^. The greater effect of surgery in older patients than in younger adults observed using the landmark analysis may be explained by the selection of the fittest patients as they had to be alive 6 months after diagnosis to be included in the analysis. Finally, we included patients with known stage IV colon cancer while 9.8% have unknown stage, which may have led to selection bias. Stage at diagnosis is more likely to be missing in older adults than younger adults^21,22^. Further studies on the effect of surgery on survival in patients diagnosed with colon cancer is, therefore, warranted.

## 7. Conclusion

Immortal time bias is often overlooked in longitudinal studies using observational data, but it is important to consider when the inclusion of participants/patients in the study does not coincide with their allocation to one of the groups being compared, as for instance, in survival analysis when comparing survival between groups defined after the start of follow-up of patients/participants (e.g., treatment). This is even more important when investigating the effect of an exposure on an outcome in different groups of individuals who may have different probabilities distributions for exposure or outcomes as in our example, where the likelihoods of early death and treatment receipt are not evenly distributed across all age groups. In all circumstances, researchers have to plan carefully their study and their analysis. For further reading on the topic we encourage interested readers to read^1,19,23,24^.

## Supporting information

R code for Main analysis

R code for simulation study

## Data Availability

CORECT-R data are available upon request at https://www.ndph.ox.ac.uk/corectr/corect-r.

## Acknowledgements

This project involves data that have been provided by, or derived from, patients and collected by the NHS as part of their care and support.

## Authors’ contributions

SP designed the study, analysed data, wrote the first version, and reviewed the manuscript. CM contributed to the design of the study and reviewed the manuscript.

CL designed and conducted the simulation study, contributed to the writing of the Methods section of the manuscript, and reviewed the manuscript.

EJAM led the CORECT-R hub, and reviewed the manuscript

## Ethics approval and consent to participate

The project is covered by the Establishing a UK Colorectal Cancer Intelligence Hub Research Ethics approval (18/SW/1034) granted by the South West – Central Bristol Research Ethics Committee on the 1^st^ June 2018. The study was performed in accordance with the Declaration of Helsinki.

This project was presented to the Bowel Cancer Intelligence UK on 30^th^ June 2021 and gained their support to progress with the work.

## Consent for publication

Not applicable

## Data availability

CORECT-R data are available upon request at https://www.ndph.ox.ac.uk/corectr/corect-r.

## References

1. Hernán MA, Sauer BC, Hernández-Díaz S, Platt R, Shrier I. Specifying a target trial prevents immortal time bias and other self-inflicted injuries in observational analyses. J Clin Epidemiol. 2016;79:70–75. DOI:10.1016/j.jclinepi.2016.04.014

2. Hanley JA, Foster BJ. Avoiding blunders involving “immortal time.” Int J Epidemiol. 2014;43(3):949–961. DOI:10.1093/ije/dyu105

3. Shariff SZ, Cuerden MS, Jain AK, Garg AX. The secret of immortal time bias in epidemiologic studies. J Am Soc Nephrol JASN. 2008;19(5):841–843. DOI:10.1681/ASN.2007121354

4. Agarwal P, Moshier E, Ru M, et al. Immortal Time Bias in Observational Studies of Time-to-Event Outcomes: Assessing Effects of Postmastectomy Radiation Therapy Using the National Cancer Database. Cancer Control J Moffitt Cancer Cent. 2018;25(1):1073274818789355. DOI:10.1177/1073274818789355

5. Ho AMH, Dion PW, Ng CSH, Karmakar MK. Understanding immortal time bias in observational cohort studies. Anaesthesia. 2013;68(2):126–130. DOI:10.1111/anae.12120

6. Pilleron S, Gower H, Janssen-Heijnen M, et al. Patterns of age disparities in colon and lung cancer survival: a systematic narrative literature review. BMJ Open. 2021;11(3):e044239. DOI:10.1136/bmjopen-2020-044239

7. Weberpals J, Jansen L, Carr PR, Hoffmeister M, Brenner H. Beta blockers and cancer prognosis - The role of immortal time bias: A systematic review and meta-analysis. Cancer Treat Rev. 2016;47:1–11. DOI:10.1016/j.ctrv.2016.04.004

8. Park HS, Gross CP, Makarov DV, Yu JB. Immortal time bias: a frequently unrecognized threat to validity in the evaluation of postoperative radiotherapy. Int J Radiat Oncol Biol Phys. 2012;83(5):1365–1373. DOI:10.1016/j.ijrobp.2011.10.025

9. van Walraven C, Davis D, Forster AJ, Wells GA. Time-dependent bias was common in survival analyses published in leading clinical journals. J Clin Epidemiol. 2004;57(7):672–682. DOI:10.1016/j.jclinepi.2003.12.008

10. Betensky RA, Mandel M. Recognizing the problem of delayed entry in time-to-event studies: Better late than never for clinical neuroscientists. Ann Neurol. 2015;78(6):839–844. DOI:10.1002/ana.24538

11. Downing A, Hall P, Birch R, et al. Data Resource Profile: The COloRECTal cancer data repository (CORECT-R). Int J Epidemiol. 2021;50(5):1418–1418k. DOI:10.1093/ije/dyab122

12. Anderson JR, Cain KC, Gelber RD. Analysis of survival by tumor response. J Clin Oncol Off J Am Soc Clin Oncol. 1983;1(11):710–719. DOI:10.1200/JCO.1983.1.11.710

13. Mi X, Hammill BG, Curtis LH, Lai ECC, Setoguchi S. Use of the landmark method to address immortal person-time bias in comparative effectiveness research: a simulation study. Stat Med. 2016;35(26):4824–4836. DOI:10.1002/sim.7019

14. Suissa S. Immortal time bias in pharmaco-epidemiology. Am J Epidemiol. 2008;167(4):492–499. DOI:10.1093/aje/kwm324

15. R Core Team. R: A Language and Environment for Statistical Computing. R Foundation for Statistical Computing; 2021. https://www.R-project.org/

16. Brilleman SL, Wolfe R, Moreno-Betancur M, Crowther MJ. Simulating Survival Data Using the simsurv R Package. J Stat Softw. 2021;97:1–27. DOI:10.18637/jss.v097.i03

17. Gouverneur A, Salvo F, Berdaï D, Moore N, Fourrier-Réglat A, Noize P. Inclusion of elderly or frail patients in randomized controlled trials of targeted therapies for the treatment of metastatic colorectal cancer: A systematic review. J Geriatr Oncol. 2018;9(1):15–23. DOI:10.1016/j.jgo.2017.08.001

18. Morris E, Treasure T. If a picture is worth a thousand words, take a good look at the picture: Survival after liver metastasectomy for colorectal cancer. Cancer Epidemiol. 2017;49:152–155. DOI:10.1016/j.canep.2017.06.009

19. Maringe C, Benitez Majano S, Exarchakou A, et al. Reflections on modern methods: trial emulation in the presence of immortal-time bias. Assessing the benefit of major surgery for elderly lung cancer patients using observational data. Int J Epidemiol. Published online May 9, 2020. DOI:10.1093/ije/dyaa057

20. Zhou Y, Abel GA, Hamilton W, et al. Diagnosis of cancer as an emergency: a critical review of current evidence. Nat Rev Clin Oncol. 2017;14(1):45–56. DOI:10.1038/nrclinonc.2016.155

21. Di Girolamo C, Walters S, Benitez Majano S, et al. Characteristics of patients with missing information on stage: a population-based study of patients diagnosed with colon, lung or breast cancer in England in 2013. BMC Cancer. 2018;18(1):492. DOI:10.1186/s12885-018-4417-3

22. Pilleron S, Charvat H, Araghi M, et al. Age disparities in stage-specific colon cancer survival across seven countries: An International Cancer Benchmarking Partnership SURVMARK-2 population-based study. Int J Cancer. Published online 2020. DOI:10.1002/ijc.33326

23. Jones M, Fowler R. Immortal time bias in observational studies of time-to-event outcomes. J Crit Care. 2016;36:195–199. DOI:10.1016/j.jcrc.2016.07.017

24. Wang J, Peduzzi P, Wininger M, Ma S. Statistical Methods for Accommodating Immortal Time: A Selective Review and Comparison. Published online February 4, 2022. Accessed February 16, 2022. https://arxiv.org/abs/2202.02369v1

