## Supplemental material for "Immortal time bias in older vs younger age groups: a simulation study with application to a population-based cohort of patients with colon cancer"

---

### List of Figures

Figure 1 – Scenarios 5-7

### List of Tables

Table 1 - Values of the shape and scale parameters of the Weibull distributions for the 7 scenarios

Table 2 – Difference in one-year overall survival based on the different methods and scenarios and its associated standard error (SE)

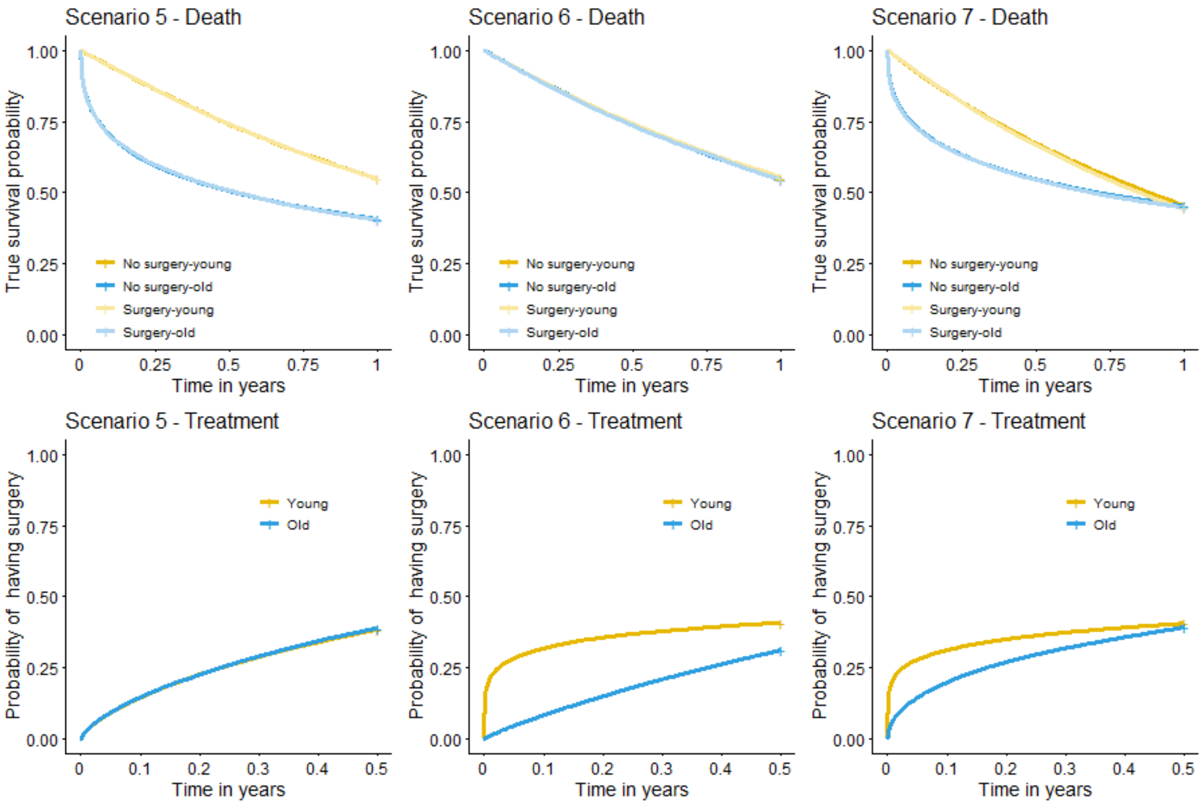


**Figure 1** – Scenarios 5-7

**Table 1** - Values of the shape and scale parameters of the Weibull distributions for the 7 scenarios

| Scenario | Treatment model | | | | Survival model | | | |
|---|---|---|---|---|---|---|---|---|
| | Younger patients | | Older patients | | Younger patients | | Older patients | |
| | Scale | Shape | Scale | Shape | Scale | Shape | Scale | Shape |
| 1 | 0,8 | 0,7 | 0,8 | 0,7 | 0,8 | 1 | 0,8 | 1 |
| 2 | 0,8 | 0,7 | 0,8 | 0,7 | 0,8 | 1 | 0,8 | 0,4 |
| 3 | 0,6 | 0,2 | 0,7 | 0,5 | 0,6 | 1 | 0,6 | 1 |
| 4 | 0,6 | 0,2 | 0,7 | 0,9 | 0,6 | 1 | 0,9 | 0,4 |
| 5 | 0,8 | 0,7 | 0,8 | 0,7 | 0,6 | 1 | 0,9 | 0,4 |
| 6 | 0,6 | 0,2 | 0,7 | 0,9 | 0,6 | 1 | 0,6 | 1 |
| 7 | 0,6 | 0,2 | 0,7 | 0,5 | 0,8 | 1 | 0,8 | 0,4 |

**Table 2** – Difference in one-year overall survival based on the different methods and scenarios and its associated standard error (SE)

| Scenario | Time-fixed Cox Model | | | | Landmark analysis | | | | Time-varying Cox model | | | | Delayed e | |
|---|---|---|---|---|---|---|---|---|---|---|---|---|---|---|
| | Older patients | | Younger patients | | Older patients | | Younger patients | | Older patients | | Younger patients | | Older patients | |
| | Estimate* | Empirical SE** | Estimate | Empirical SE | Estimate | Empirical SE | Estimate | Empirical SE | Estimate | Empirical SE | Estimate | Empirical SE | Estimate | Empirical SE |
| 1 | -0.133 | 0.045 | -0.134 | 0.043 | 0.000 | 0.048 | -0.007 | 0.054 | -0.001 | 0.049 | -0.005 | 0.047 | 0.000 | 0.007 |
| 2 | -0.302 | 0.045 | -0.134 | 0.043 | 0.001 | 0.046 | -0.007 | 0.054 | -0.005 | 0.061 | -0.004 | 0.047 | -0.001 | 0.005 |
| 3 | -0.092 | 0.044 | -0.049 | 0.041 | -0.003 | 0.042 | -0.007 | 0.047 | 0.000 | 0.047 | -0.006 | 0.042 | 0.000 | 0.009 |
| 4 | -0.319 | 0.047 | -0.049 | 0.041 | 0.001 | 0.053 | -0.007 | 0.047 | 0.003 | 0.063 | -0.006 | 0.042 | 0.002 | 0.005 |
| 5 | -0.311 | 0.042 | -0.118 | 0.043 | 0.006 | 0.052 | -0.001 | 0.044 | 0.004 | 0.059 | -0.002 | 0.049 | 0.000 | 0.005 |
| 6 | -0.120 | 0.045 | -0.048 | 0.044 | -0.002 | 0.051 | -0.001 | 0.043 | 0.008 | 0.052 | -0.004 | 0.046 | 0.004 | 0.012 |
| 7 | -0.255 | 0.045 | -0.052 | 0.044 | 0.006 | 0.047 | 0.001 | 0.055 | 0.008 | 0.058 | -0.001 | 0.045 | 0.000 | 0.003 |

* Average estimated difference in 1-year overall survival probabilities across the 1000 simulations. The true value is 0.
** Standard deviation of the 1000 estimated difference in 1-year overall survival probabilities
