## Supplementary material for "Immortal time bias in older vs younger age groups: a simulation study with application to a population-based cohort of patients with colon cancer": R code for simulation study

#######################################
# Simulations immortal-time bias
# Author: C. leyrat
# Date: 08/05/2021
# Input: none
# Output: results.csv
#######################################

#packages
library(survival) 
library(simsurv)
library(boot)


###############################################################################

########################
# Data generation
######################## 

generation<-function(n,p_age,vect_param){
  
  #Generation of the patients' ID and covariates (only binary age to start with)
    covs <- data.frame(id = 1:n, age = rbinom(n, 1, p_age))
  
  #Splitting it by age
    covs_y <- covs[covs$age==0,]
    covs_o <- covs[covs$age==1,]
  
  #Scale and shape parameters for the treatment distribution
  #Among younger patients
    scale_trt_y<-vect_param[1]  
    shape_trt_y<-vect_param[2]  
  
  #Among older patients
    scale_trt_o<-vect_param[3]  
    shape_trt_o<-vect_param[4] 
  
  
  #Scale and shape parameters for the survival distribution
  #Among younger patients
    scale_death_y<-vect_param[5]
    shape_death_y<-vect_param[6]
  
  #Among older patients
    scale_death_o<-vect_param[7]
    shape_death_o<-vect_param[8]
  
  
  ##############################################################################
  #Simulation of treatment status and time-to-treatment (Weibull distribution)
  ##############################################################################
  
    #Different shape per age group
    # Maximum follow-up time is 6 months (maxt)
    
    #Among the young
      s1_y <- simsurv(lambdas = scale_trt_y, gammas = shape_trt_y, 
                      x = covs_y, maxt = 0.5)
    
    #simsurv changes the ids, so we manually get the initial patients ids
      s1_y$id<-covs_y$id
    
    
    #Among the older patients
      s1_o <- simsurv(lambdas = scale_trt_o, gammas = shape_trt_o, 
                      x = covs_o, maxt = 0.5)
      s1_o$id<-covs_o$id
    
    #Combining the two datasets
      s1<-rbind(s1_y,s1_o)
    
    #Rename the variables
      names(s1)<-c("id","time_trt_true","trt_true")
      dat<-merge(covs,s1, by="id")
  
      
  #######################################################################
  #Simulation of survival status and time-to-event (Weibull distribution)
  #######################################################################
      
  # Maximum follow-up time is 1 year (maxt)
  #Among the young
    s2_y <- simsurv(lambdas = scale_death_y, gammas = shape_death_y, 
                    x = covs_y, maxt = 1)
    s2_y$id<-covs_y$id
  
  #Among the older patients
    s2_o <- simsurv(lambdas = scale_death_o, gammas = shape_death_o, 
                    x = covs_o, maxt = 1)
    s2_o$id<-covs_o$id
  
    #Combining the two datasets
    s2<-rbind(s2_y,s2_o)
    
  #Rename the variables
    names(s2)<-c("id","time_death","death")
    dat<-merge(dat,s2, by="id")
  
    
  #Find patients whose time-to-surgery is longer than time-to-death
  #This is where we introduce the immortal-time bias
    dat$order<-ifelse(dat$trt_true==1 & dat$time_trt>dat$time_death,0,1)
  
  #Creation of the observed treatment variable and time-to-treatment
    dat$trt_obs<-ifelse(dat$order==0,0,dat$trt_true)
    dat$time_trt_obs<-ifelse(dat$order==0,dat$time_death,dat$time_trt_true)
    
  #returning the dataset for the analysis
    return(dat)
    
}


#####################################################################################################

#####################################
# Descriptive statistics
#####################################

desc<-function(dat){
  
  #True proportion of treated per age group
    prop_trt_y_true<-mean(dat$trt_true[dat$age==0])
    prop_trt_o_true<-mean(dat$trt_true[dat$age==1])
    
  #Observed proportion of treated per age group
    prop_trt_y_obs<-mean(dat$trt_obs[dat$age==0])
    prop_trt_o_obs<-mean(dat$trt_obs[dat$age==1])
  
  #1 year survival probabilities per treatment group (true treatment) and age group
  surv1<-survfit(Surv(time_death, death) ~ trt_true+age, data = dat)
  psurv_true<-as.data.frame(t(summary(surv1,time=1)$surv))
  names(psurv_true)<-c("S0_y_true","S0_o_true","S1_y_true","S1_o_true")
  
  #1 year survival probabilities per treatment group (observed treatment)
  surv2<-survfit(Surv(time_death, death) ~ trt_obs+age, data = dat)
  psurv_obs<-as.data.frame(t(summary(surv2,time=1)$surv))
  names(psurv_obs)<-c("S0_y_obs","S0_o_obs","S1_y_obs","S1_o_obs")
  
  res<-cbind(prop_trt_y_true,prop_trt_o_true,prop_trt_y_obs,prop_trt_o_obs,psurv_true,psurv_obs)
  
  return(res)
}


########################
#Analysis
########################


##############################
# Method 1: naive Cox model
##############################

naive_cox<-function(dat){

 
  #2 Cox by age group
    mcox_y<-coxph(Surv(time_death, death)~trt_obs, ties="efron", data=dat[dat$age==0,])
    mcox_o<-coxph(Surv(time_death, death)~trt_obs, ties="efron", data=dat[dat$age==1,])
    
  #Creation of 2 datasets: one with trt_obs=0 and one with trt_obs=1
  #Note: this would be necessary to obtain marginal effect if we had other covariates in the model,
  #but without covariates we could just take the mean predictions among the subgroup of interests
    d0_y<-d1_y<-dat[dat$age==0,]
    d0_o<-d1_o<-dat[dat$age==1,]
    
  #We want to predict at 1 year as if there was no censoring, so we chnge the values in the new datasets
    d0_y$death<-d1_y$death<-d0_o$death<-d1_o$death<-1
    d0_y$time_death<-d1_y$time_death<-d0_o$time_death<-d1_o$time_death<-1
    d0_y$trt_obs<-d0_o$trt_obs<-0
    d1_y$trt_obs<-d1_o$trt_obs<-1
    
  #Now we can obtain the predictions, as if everyone was treated vs not
    d0_y$pred<-exp(-predict(mcox_y, newdata=d0_y, type="expected"))
    d1_y$pred<-exp(-predict(mcox_y, newdata=d1_y, type="expected"))
    
    d0_o$pred<-exp(-predict(mcox_o, newdata=d0_o, type="expected"))
    d1_o$pred<-exp(-predict(mcox_o, newdata=d1_o, type="expected"))
    
  # Calculationg the differences in one year survival from the predictions

  #Among the younger patients
    diff_surv_cox_young<-mean(d0_y$pred[d0_y$age==0])-mean(d1_y$pred[d1_y$age==0])
    
  #Among the older patients
    diff_surv_cox_old<-mean(d0_o$pred[d0_o$age==1])-mean(d1_o$pred[d1_o$age==1])
    
  #Concatenating the results
    res2<-c(diff_surv_cox_young,diff_surv_cox_old)
    names(res2)<-c("diff_surv_cox_young","diff_surv_cox_old")

    return(res2)
}


###################################
# Method 2: time-varying Cox model
###################################

  timevar_cox<-function(dat){
    
  #Vector of times of events in the whole dataset (death)
    t_events<-sort(unique(dat$time_death))
    times<-data.frame("tevent"=t_events,"ID_t"=seq(1:length(t_events)))
    
  #Creation of the entry variable (Tstart, 0 for everyone)
    dat$Tstart<-0
    
  #Splitting the dataset at each time of event
   #until the event happens (id will be a new variable identifying the patient)
    data.long<-survSplit(dat, cut=t_events, end="time_death", 
                         start="Tstart", event="death",id="id_pat") 
    
    data.long$trt_varying<-ifelse(data.long$trt_obs==1 & data.long$Tstart>=data.long$time_trt_obs,1,0)
    
 
  #2 Cox by age group
    mcox_var_y<-coxph(Surv(Tstart,time_death, death)~trt_varying, ties="efron", 
                      data=data.long[data.long$age==0,],
                    control = coxph.control(timefix = FALSE))
    mcox_var_o<-coxph(Surv(Tstart,time_death, death)~trt_varying, ties="efron", 
                      data=data.long[data.long$age==1,],
                      control = coxph.control(timefix = FALSE))
    
  #Creation of 2 datasets: one with trt_obs=0 and one with trt_obs=1
  #Note: this would be necessary to obtain marginal effect if we had other covariates in the model,
  #but without covariates we could just take the mean predictions among the subgroup of interests
    d0_y<-d1_y<-dat[dat$age==0,]
    d0_o<-d1_o<-dat[dat$age==1,]
    
  #We want to predict at 1 year as if there was no censoring, so we chnge the values in the new datasets
    d0_y$death<-d1_y$death<-d0_o$death<-d1_o$death<-1
    d0_y$time_death<-d1_y$time_death<-d0_o$time_death<-d1_o$time_death<-1
    d0_y$trt_varying<-d0_o$trt_varying<-0
    d1_y$trt_varying<-d1_o$trt_varying<-1
    
  #Now we can obtain the predictions, as if everyone was treated vs not
    d0_y$pred<-exp(-predict(mcox_var_y, newdata=d0_y, type="expected"))
    d1_y$pred<-exp(-predict(mcox_var_y, newdata=d1_y, type="expected"))
    
    d0_o$pred<-exp(-predict(mcox_var_o, newdata=d0_o, type="expected"))
    d1_o$pred<-exp(-predict(mcox_var_o, newdata=d1_o, type="expected"))
    
  # Calculationg the differences in one year survival from the predictions
    
  #Among the younger patients
    diff_surv_cox_young<-mean(d0_y$pred[d0_y$age==0])-mean(d1_y$pred[d1_y$age==0])
    
  #Among the older patients
    diff_surv_cox_old<-mean(d0_o$pred[d0_o$age==1])-mean(d1_o$pred[d1_o$age==1])
    
  #Concatenating the results
    res2<-c(diff_surv_cox_young,diff_surv_cox_old)
    names(res2)<-c("diff_surv_timevar_young","diff_surv_timevar_old")
    
    return(res2)
  }


 
  ##############################
  # Method 3: landmark analysis
  ##############################
  
  landmark_cox<-function(dat){
    
    #Selecting only patients alive at 6 months
      dat<-dat[dat$time_death>=0.5,]
    
    
    #2 Cox by age group
      mcox_y<-coxph(Surv(time_death, death)~trt_obs, ties="efron", data=dat[dat$age==0,])
      mcox_o<-coxph(Surv(time_death, death)~trt_obs, ties="efron", data=dat[dat$age==1,])
    
    #Creation of 2 datasets: one with trt_obs=0 and one with trt_obs=1
    #Note: this would be necessary to obtain marginal effect if we had other covariates in the model,
    #but without covariates we could just take the mean predictions among the subgroup of interests
      d0_y<-d1_y<-dat[dat$age==0,]
      d0_o<-d1_o<-dat[dat$age==1,]
    
    #We want to predict at 1 year as if there was no censoring, so we chnge the values in the new datasets
      d0_y$death<-d1_y$death<-d0_o$death<-d1_o$death<-1
      d0_y$time_death<-d1_y$time_death<-d0_o$time_death<-d1_o$time_death<-1
      d0_y$trt_obs<-d0_o$trt_obs<-0
      d1_y$trt_obs<-d1_o$trt_obs<-1
    
    #Now we can obtain the predictions, as if everyone was treated vs not
      d0_y$pred<-exp(-predict(mcox_y, newdata=d0_y, type="expected"))
      d1_y$pred<-exp(-predict(mcox_y, newdata=d1_y, type="expected"))
      
      d0_o$pred<-exp(-predict(mcox_o, newdata=d0_o, type="expected"))
      d1_o$pred<-exp(-predict(mcox_o, newdata=d1_o, type="expected"))
    
    # Calculationg the differences in one year survival from the predictions
    
    #Among the younger patients
      diff_surv_cox_young<-mean(d0_y$pred[d0_y$age==0])-mean(d1_y$pred[d1_y$age==0])
    
    #Among the older patients
      diff_surv_cox_old<-mean(d0_o$pred[d0_o$age==1])-mean(d1_o$pred[d1_o$age==1])
    
    #Concatenating the results
      res2<-c(diff_surv_cox_young,diff_surv_cox_old)
      names(res2)<-c("diff_surv_landmark_young","diff_surv_landmark_old")
      
      return(res2)
  }
 


  ######################################
  # Method 4: delayed entry
  ######################################
  
  delayed<-function(dat){
    

    ##############
    # By subgroup
    ##############
    
    #First model among treated patients only
      dat$var<-1
      dat1<-dat[dat$trt_obs==1,]
      dat1$exit<-dat1$time_death-dat1$time_trt_obs
      dat1$event<-dat1$death
    
      dat1_y<-dat1[dat1$age==0,]
      dat1_o<-dat1[dat1$age==1,]
    
    #One model per age group
      m1_y<-coxph(Surv(exit,event)~1, ties="efron", data=dat1_y)
      m1_o<-coxph(Surv(exit,event)~1, ties="efron", data=dat1_o)
     
    #Second model among untreated patients and treated patients before treatment 
      dat0<-dat
      dat0$exit<-ifelse(dat0$trt_obs==0,dat0$time_death,dat0$time_trt_obs)
      dat0$event<-ifelse(dat0$trt_obs==0,dat0$death,0)
      
      dat0_y<-dat1[dat0$age==0,]
      dat0_o<-dat1[dat0$age==1,]
    
    #One model per age group
      m0_y<-coxph(Surv(exit,event)~1, ties="efron", data=dat0_y)
      m0_o<-coxph(Surv(exit,event)~1, ties="efron", data=dat0_o)
      
      d<-dat
      d$S1<-ifelse(d$age==0,exp(-max(basehaz(m1_y)[1,])),exp(-max(basehaz(m1_o)[1,])))
      d$S0<-ifelse(d$age==0,exp(-max(basehaz(m0_y)[1,])),exp(-max(basehaz(m0_o)[1,])))
    
    # Calculationg the differences in one year survival from the predictions
    #Among the younger patients
      diff_surv_cox_young<-mean(d$S0[d$age==0])-mean(d$S1[d$age==0])
    
    #Among the older patients
      diff_surv_cox_old<-mean(d$S0[d$age==1])-mean(d$S1[d$age==1])
    
    #Concatenating the results
      res2<-c(diff_surv_cox_young,diff_surv_cox_old)
      names(res2)<-c("diff_surv_delayed_young","diff_surv_delayed_old")
      
      return(res2)
  }
  

  
  #################################################################
  # Analysis function for the bootstrap
  # Note: time-varying Cox not included because of computing time
  #################################################################
  
  analysis_boot<-function(dat,indices){
  
  #Selection of bootstrap sample  
    d<-dat[indices,]
    
  #Conducting the analysis
    naive<-naive_cox(d)
    landmark<-landmark_cox(d)
    delay<-delayed(d)
    timevar<-timevar_cox(d)
    res<-c(naive,landmark,timevar,delay) 
    return (res)
  }
  


  #############################
  # Bootstrap implementation
  #############################
  
  fboot<-function(dat,R){
    
    #Extracting the names of the different estimates for later
      method<-names(analysis_boot(dat,1:1000))
    
    #Running R bootstraps
      b<-boot(dat, analysis_boot, R=R)

    #Extracting the initial estimates and bootsrap SE (and Var)
      res_boot<-data.frame("Estimate"=b$t0, "SE"=apply(b$t, 2, sd), "Var"=(apply(b$t, 2, sd))^2)
    
    #Constructing a normal-based 95%CI
      res_boot$low<-res_boot$Estimate-qnorm(0.975)*res_boot$SE
      res_boot$up<-res_boot$Estimate+qnorm(0.975)*res_boot$SE
    
    #Indicator of whether the CI contains 0 (for the coverage rate)
      res_boot$sig<-ifelse(res_boot$low<0 & res_boot$up>0, 0, 1)
    
    #Adding a variable wih the name of the methods
      res_boot$method<-method
      
      return(res_boot)
  }


  ###########################################
  # Final simulation function
  ###########################################
  
  sim<-function(n,p_age,vect, R, nsim){
    
    #Setting the RNG seed
     # set.seed(1604*IDtask)
     # Sys.sleep(IDtask*90)
    
    descriptive<-NULL
    results<-NULL

    #Generating and analysing nsim dataset
      for (i in (1:nsim)){
        print(i)
        dat<-generation(n,p_age,vect) 
        descr<-desc(dat)
        analysis<-fboot(dat,R)
        timevar<-timevar_cox(dat)
        descriptive<-rbind(descriptive, descr)
        results<-rbind(results, analysis)
      }
    
    #Averaging the results over the nsim simulations
      summary_desc<-colMeans(descriptive)
    
    #Calculating relevant measures (empirical SE and MCE)
      t<-results
      summary<-aggregate(t[, c(1,3:6)], list(t$method), mean)
      summary$mod_SE<-sqrt(summary$Var)
      summary$var_var<-(aggregate(t[, c(3)], list(t$method), sd)[,2])^2
      summary$emp_var<-(aggregate(t[, c(1)], list(t$method), sd)[,2])^2
      summary$MCE_Estimate<-sqrt(summary$emp_var/nsim)
      summary$MCE_emp_SD<-sqrt(summary$emp_var/(2*(nsim-1)))
      summary$MCE_mod_SE<-sqrt(summary$var_var/(4*(nsim*summary$Var)))
      summary$MCE_coverage<-sqrt(summary$sig*(1-summary$sig)/nsim)
      
      #summary$scenario<-IDtask
     
      #name<-paste("descriptive_",IDtask,".csv",sep="")
      #name2<-paste("summary_",IDtask,".csv",sep="")
      #name3<-paste("descriptive_tot_",IDtask,".csv",sep="")
      #name4<-paste("summary_tot_",IDtask,".csv",sep="")
      #write.csv(summary_desc,name,row.names=F)
      #write.csv(summary,name2,row.names=F)
      #write.csv(descriptive,name3,row.names=F)
      #write.csv(results,name4,row.names=F)
      
    #Return the full results + aggregated results 
    return(list(descriptive,results,summary_desc, summary))
  }
  

  #Example with only 3 repetitions and 10 bootstrap replicates
  a<-sim(1000,0.5,c(0.8,0.7,0.8,0.7,0.8,1,0.8,1),10,3)
  a
  

 

  
  
